# High Throughput Screening for Expanded CTG repeats in Myotonic Dystrophy Type 1 Using Melt Curve Analysis

**DOI:** 10.1101/2021.01.11.21249609

**Authors:** Russell J Butterfield, Carina Imburgia, Katie Mayne, Tara Newcomb, Diane M Dunn, Brett Duval, Marcia L Feldkamp, Nicholas E Johnson, Robert B Weiss

## Abstract

**Background:** Myotonic dystrophy type 1 (DM1) is caused by CTG repeat expansions in the *DMPK* gene and is the most common form of muscular dystrophy. Patients can have long delays from onset to diagnosis, since clinical signs and symptoms are often non-specific and overlapping with other disorders. Clinical genetic testing by Southern blot or triplet-primed PCR (TP-PCR) is technically challenging and cost prohibitive for population surveys.

**Methods:** Here, we present a high throughput, low-cost screening tool for CTG repeat expansions using TP-PCR followed by high resolution melt curve analysis with saturating concentrations of SYBR GreenER dye.

**Results:** We determined that multimodal melt profiles from the TP-PCR assay are a proxy for amplicon length stoichiometry. In a screen of 10,097 newborn blood spots, melt profile analysis accurately reflected the tri-modal distribution of common alleles from 5 to 35 CTG repeats, and identified the premutation and full expansion alleles.

**Conclusion:** We demonstrate that robust detection of expanded CTG repeats in a single tube can be achieved from samples derived from specimens with minimal template DNA such as dried blood spots (DBS). This technique is readily adaptable to large-scale testing programs such as population studies and newborn screening programs.

## 1 INTRODUCTION

Myotonic dystrophy type 1 (DM1) is an autosomal dominant disorder characterized by early onset cataracts, myotonia, muscle weakness, cardiac arrhythmias, respiratory failure, and gastrointestinal dysfunction.^1^ DM1 is caused by an expansion of the trinucleotide CTG repeat in the 3’ non-coding region of the *DMPK* gene (OMIM #605377) on chromosome 19q13.3.^2-4^ Expansions of ≥50 CTG repeats are considered pathogenic, while expansions of 35-49 CTG repeats result in a premutation state at risk for passing expanded alleles to the next generation. The mutational reservoir for most expanded alleles is an ancient linkage disequilibrium block with 18-35 CTG repeat alleles that varies in haplotype frequency between human populations.^5^

DM1 is the most common form of muscular dystrophy with prevalence estimates between 0.5 and 1.5 per 10,000.^6-8^ Diagnosis is based on clinical signs and symptoms that are often non-specific and overlapping with other disorders, followed by genetic testing to detect the CTG repeat expansion in *DMPK*. Given the complexity of diagnosis and variability of clinical symptoms, diagnostic delays of 7 years or more from onset of symptoms are common.^9^ Diagnosis in muscular dystrophies, even in very rare forms of muscular dystrophy has been simplified by the proliferation of next-generation sequencing panels and exome sequencing, however, triplet repeat disorders such as DM1 are not routinely identified by these sequencing methods, and many patients with DM1 continue to have long diagnostic delays.

The gold standard for clinical genetic testing for DM1 uses a Southern blot technique to estimate the size of the CTG expansion.^10^ There are a number of challenges with this technique, including the requirement for a large amount of DNA and the amount of time required to perform the method. More recently, fluorescently labeled triplet-primed PCR (TP-PCR) and fragment sizing by capillary electrophoresis has been proposed as an alternative for diagnosis of DM1^11, 12^ and for other triplet repeat expansion disorders such as Fragile X syndrome and Huntington disease^13, 14^. The TP-PCR methodology uses a (CTG)_n_ or (CAG)_n_ repeat primer to generate a nested set of fragments from the expanded allele, and the observed ‘ladder’ of amplicons is detected by capillary electrophoresis. Both Southern blot and TP-PCR with capillary electrophoresis are technically challenging and limited by cost and difficulty of performing the assay on a large scale, such as in a population screen. A simplified approach using TP-PCR combined with a melt curve analysis (MCA) has been developed for detection of repeat expansions in several triplet repeat disorders including DM1, Fragile X syndrome, and Huntington disease.^15-17^ This TP-PCR with MCA approach in DM1 is based on nonsaturating SYBR Green I intercalating dye to detect expansion of the (CTG)_n_ repeat in *DMPK*.^16^ In this technique, a slow temperature ramp allows the limiting SYBR Green I dye to re-equilibrate to longer duplexes, thus suppressing the melt detection of shorter, lower *T*_m_ amplicons while maintaining the ability to detect decreased fluorescence from the melting of larger, higher *T*_m_ amplicons. Under conditions optimized for the balance between dye and amplicon concentration, this technique preferentially detects higher length products in the nested set of TP-PCR fragments, reducing a multimodal melt curve to a unimodal profile. The requirement for balancing the limiting dye concentration versus the concentration of the longer TP-PCR fragments may be challenging for crude DNA inputs lower than 5 ng, such as might be obtained from a dried blood spot (DBS) sample commonly used in population screens.

Saturating intercalating dyes such as SYBR GreenER, EvaGreen, and LCGreen have been developed for high-resolution melting analysis of multimodal melt curves. These dyes are less inhibitory to PCR and can be used at a higher concentration to reduce the level of dye relocation during melting. Here, we demonstrate that by using SYBR GreenER dye during TP-PCR with MCA, we can achieve accurate detection of expanded CTG repeats from DBS samples using the full multimodal melt profiles from the nested set of amplicons. This technique uses a simplified DNA extraction from a 3mm DBS sample and provides a rapid, inexpensive test for expansion of the (CTG)_n_ repeat in the *DMPK* gene that is readily adaptable to large-scale screening programs such as population studies and newborn screening programs.

## 2 METHODS

### 2.1 Ethical Compliance

The study was performed under University of Utah Institutional Review Board approved protocols (IRB #40092 and IRB #87466).

### 2.2 Study Samples

Genomic DNA for control samples was purified from whole blood from individuals with a clinical diagnosis of DM1 and from unaffected controls after providing written informed consent. Control samples included fully expanded, clinically affected subjects with DM1 (≥50 CTG repeats), subjects with premutation at high risk for passing expanded repeats in progeny (35-49 CTG repeats), subjects with intermediate expansions of the CTG repeat but still in the normal range (18-34 CTG repeats), and normal subjects with 5-17 CTG repeats. Genomic DNA for the haplotype analysis was derived from non-DM1 individuals with high density single nucleotide polymorphism (SNP) genotypes to identify inheritance of the three common *DMPK* haplotypes.^5^

Population based test samples were derived from 10,224 consecutive anonymous newborn DBS from the Newborn Screening Program at the New York State Newborn Screening Program, Wadsworth Center, New York State Department of Health. The DBS did not include samples known to be repeats from newborns already sampled, those whose parents requested that specimens not be used for research, those that were unsuitable for screening/DNA analysis, and specimens for which there wasn’t enough blood left for research. Three-millimeter punches were individually placed into 96-well non-skirted polypropylene PCR plates with each plate containing nine consecutive blank spaces for control samples. The control row was rotated from row A to H and after shipment to Utah, plates were stored at 4 ^°^C until DNA purification was performed.

### 2.3 DNA Purification From Dried Blood Spots

We modified the low-cost CASM method developed at the New York State Dept. of Health to extract DNA from DBS ^18^. Four 96-well plates were processed in batches beginning with the addition of 150 μl per well of Red Blood Cell (RBC) lysis solution (10 mM Tris-HCl, 320 mM Sucrose, 2.4 mM MgCl_2_, 1% Triton X-100, pH 8.0). The plates were covered with clear acetate plate sealers, briefly centrifuged and shaken for 20 min at 750 rpm on a Heidolph Titramax 1000 plate shaker. Subsequent reagent additions were done with a Velocity11 VPrep 96-channel pipetting station and liquid removal was done manually with 12-channel pipettors. After removal of the RBC lysis solution, the DBS were washed twice with 160 μl of dH_2_O and shaken for 30 min. After this, 50 μl of 100 mM NaOH, 2% Tween 20 was added per well, the plates were sealed and heated to 95 ^°^ C for 12 min. After briefly centrifuging the plates, 50 μl of 100 mM Tris-HCl, 2 mM EDTA, pH 8.0 was added per well, the plates were shaken for 10 min, briefly centrifuged and the lysate containing genomic DNA was transferred to a fresh 96-well plate (0.2 ml non-skirted Thermo Scientific PCR plate), covered with foil sealers and stored at 4^°^ C. The concentration of genomic DNA isolated from 939 DBS was estimated from real-time PCR comparing cycle threshold (Ct) values to known concentrations using a linear model with a median of 1.11 ng/µl (lower quartile 0.15ng/µl and upper quartile 6.44 ng/µl).

### 2.4 Triplet Primed PCR (TP-PCR) and Melt Curve Analysis (MCA)

TP-PCR for both 5’ and 3’ ends of the (CTG)_n_ repeat in *DMPK* (NM_004409.5) were adapted from conditions published by Falk et al.^19^ and optimized for use of SYBR® GreenER™ dye to facilitate MCA (**Figure 1A**). For the 5’ TP-PCR, amplification was performed in 384-well PCR plates with 10 μl reaction volumes consisting of 1 μl (∼1ng) genomic DNA and the following 9 μl reagent master mix: 0.5 μM *DMPK* forward primer (5’-GGGGCTCGAAGGGTCCTTGT-3’), 0.05 μM (CAG)_6_ reverse primer (5’-AGCGGATAACAATTTCACACAGGACAGCAGCAGCAGCAGCAG-3’), 0.5 μM tail primer (5’-AGCGGATAACAATTTCACACAGGA-3’), 0.5 M GC-RICH Resolution Solution (Roche), and 1X SYBR Select Master Mix (Thermo Fisher Scientific, cat. # 4472913, contains SYBR GreenER dye, AmpliTaq DNA Polymerase UP, dNTPs with dUTP/dTTP, heat-labile UDG, ROX passive reference dye and buffer components). For 3’ TP-PCR, amplification was performed using the same procedure but substituting 0.5 μM *DMPK* reverse primer (5’-GTGCGTGGAGGATGGAAC-3’), 0.05 μM (CTG)_6_-forward primer (5’-AGCGGATAACAATTTCACACAGGATGCTGCTGCTGCTGCTGCTG-3’), and 0.5 μM tail primer (5’-AGCGGATAACAATTTCACACAGGA-3’). Thermocycling was performed on either a MJ Research PTC-225 or an Applied Biosystems Veriti 384-well Thermal Cycler with the following cycling parameters: UDG activation of 50 ^°^C for 2 min followed by AmpliTaq activation at 95 ^°^C for 1 min, followed by 50 cycles of denaturation at 95 ^°^C for 1 min, annealing at 60 ^°^C for 1 min, and extension at 72^°^C for 3 min. A 72 ^°^C final extension for 10 min followed by a 4 ^°^C hold completed the reaction; plates were stored at -20 ^°^C. After PCR cycling and storage, samples underwent high resolution MCA on an Applied Biosystems QuantStudio 12K Flex instrument with a single cycle of 98 ^°^C for 15 seconds, 60 ^°^C for 15 seconds, and final ramp to 98 ^°^C at 0.05 ^°^C /s. Temperature, normalized fluorescence intensity (Rn) and the negative derivative of Rn with respect to temperature (-dF/dT) were exported using the QuantStudio 12K Flex software. For screens using DBS samples, two sets of the nine TP-PCR amplification and MCA controls were included in each 384-well plate. The nine control samples, with CTG repeat size is denoted as allele1::allele 2, included four in the normal/intermediate range (5::5, 5::13, 5::14, and 14::30), one in the premutation range (12::37), and four in the fully expanded range (5::75, 5::80, 5::480, and 14::2530).

**Figure 1.**
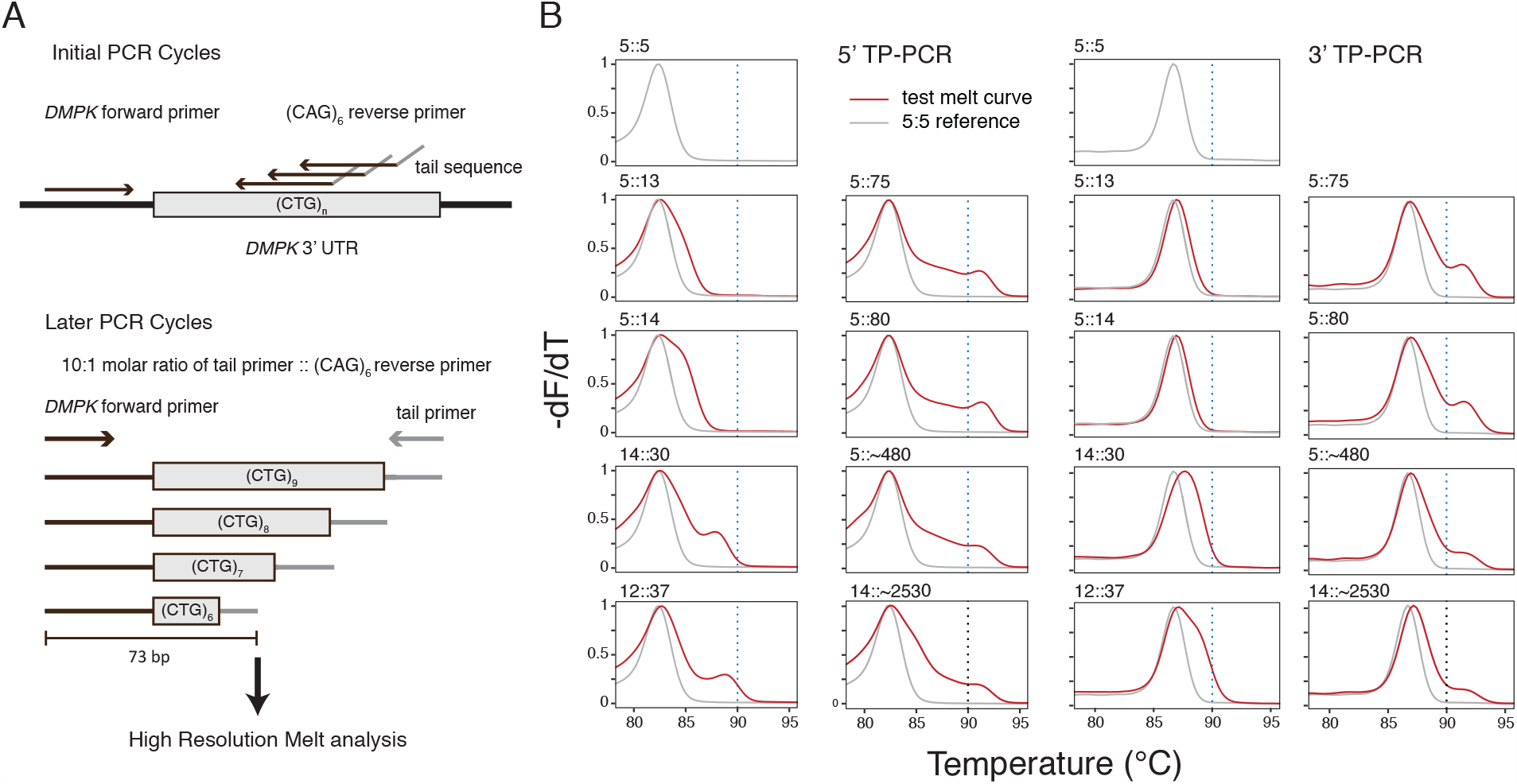
Assay design for *DMPK* TP-PCR melt curve analysis. Location of the unique *DMPK* forward primer and the (CAG)_6_ reverse / tail primers used to generate a nested set of amplicons. (B) Normalized -dF/dT melt curves from the 5’ TP-PCR and 3’ TP-PCR reactions using control samples with increasing CTG repeat lengths. The melt curve from the 5:5 homozygous sample (gray curve) is plotted with melt curve from each test sample (red curve). The CTG repeat sizes of the alleles in the test samples are indicated above each plot.

### 2.5 Primer Extension and Melt Curve Reconstruction

Primer extension reactions on post-PCR amplicons were used to reconstruct predicted melt curve profiles. PCR primers and nucleotides were removed after TP-PCR amplification with ExoSAP-IT (Thermo Fisher Scientific) and 10 μl primer extension reactions were composed of: 10 mM Tris-HCl, pH 8.3, 50 mM KCl, 1.5 mM MgCl_2_, 0.2 mM dNTPs, 0.05 μl Platinum *Taq* DNA polymerase with 0.36 μl of KB Extender (Thermo Fisher Scientific), 2 pM of 5’ 6-FAM-labeled *DMPK* forward primer, and 2 μl of ExoSAP-IT treated post-PCR amplicons. Thermocycling conditions included an initial denaturation at 95 °C for 5 min., followed by 10 cycles of 95 °C for 15 sec, 55 °C for 15 sec, 68 °C for 30 sec and a final extension at 68 °C for 7 min. 3 μl of the primer extension reactions were added to 10 μl of Hi-Di Formamide containing 0.5 μl of GeneScan 500 LIZ dye size standard and fragment analysis was performed on an ABI3730 capillary electrophoresis instrument. Electropherograms were processed using R library functions in the *seqinr* package to assign fragment sizes and peak heights.^20^ The melt curve reconstruction used a series of TP-PCR fragment sequences beginning with 5’-GGGGCTCGAAGGGTCCTTGTAGCCGGGAATG(CTG)_n_AAATTGTTATCCGCT-3’, where n ranged from 5 to 88 CTG repeats. The prediction of -d(Helicity)/d(Temp) for each fragment size was modeled by the uMelt Batch software using the default (SantaLucia) thermodynamic library with parameters adjusted to: [Mono^+^] = 22 mM, free [Mg^++^] = 1.7 mM, and DMSO = 8%.^21^ Predictions were in the temperature range of 60-98^°^C at 0.1^°^C resolution and the fragment size ratios seen by primer extension were used to generate a composite melt curve.

### 2.6 CTG Repeat Size Confirmation by Fragment Analysis

Genomic DNA selected for direct sizing analysis was further purified using Zymo DNA Clean and Concentrator-5 columns (Zymo Research, Irvine, CA) in order concentrate the sample for optimal use in follow-up assays. Fifty μl of DNA derived from the DBS was purified according to the manufacturer’s specifications and samples were eluted in 25 μl. Direct PCR amplification of the (CTG)_n_ repeat from 2 μl of Zymo purified DNA was performed in 25 μl reactions composed of: 10 mM Tris-HCl, pH 8.3, 50 mM KCl, 1.5 mM MgCl_2_, 0.2 mM dNTPs, 0.75 μl of KB Extender, 0.1 μl Platinum *Taq* DNA polymerase (Invitrogen, cat. #10966-034), 5 pM of primers (6-FAM-labeled *DMPK* forward primer and *DMPK* reverse primer: 5’-GTGCGTGGAGGATGGAAC-3’). Thermocycler conditions included an initial denaturation at 95 °C for 5 min followed by 33 cycles of 95 °C for 15 sec, 55 °C for 15 sec, 68 °C for 30 sec and a final extension at 68 °C for 7min. Fragment analysis was performed on an ABI3730xl capillary instrument and electropherograms were processed as described above for primer extensions. Samples suspected to have expansions of >75 CTG repeats were further validated using the AmplideX PCR/CE *DMPK* Kit (Asuragen, Austin, TX) and PCR reactions were carried out according to the manufacturer’s specifications. 2 μl of the amplicons were added to 10 μl of Hi-Di formamide containing 2 μl of ROX 1000 size ladder (Asuragen, Austin, TX) and electrophoresed on an ABI3730xl instrument. The electropherograms were analyzed using GeneMapper v.4.0 and repeat sizes were determined using the Asuragen Macrotable software.

### 2.7 Classification of Melt Curve Profile and Clustering

Derivative plots of normalized -dF/dT values for each sample were plotted from 80 °C to 94 °C with averaged -dF/dT melt curves from 4 positive controls with full expansion (≥50 CTG repeats) and a vertical threshold at 90 °C as reference guides for scoring (**Figure S1**). Individual plots for each sample were scored in a blinded fashion by four reviewers and classified into one of six categories based on visual inspection of the melt curve profile: normal, intermediate, premutation, expanded, uncertain, and fail. A training set of melt profiles with known CTG repeat sizes was used to establish after group review, a consensus for the criteria used to assign a melt profile to a category. Final classification for each sample was made by the majority call from the four blinded reviewers. In the case of a tie, the higher (expanded > premutation >intermediate > normal) classification was assigned. For unsupervised clustering analysis, normalized -dF/dT data interpolated to 0.05 ^°^C increments and a temperature range of 87.25 ^°^C-96.70 ^°^C was used for clustering with the Uniform Manifold Approximation and Projection (*UMAP*) dimension reduction technique and visualized with the fit_transform method and parameters of min_dist = 0.6, n_neighbors = 200, and n_components = 3.^22^

### 2.8 Haplotype/Diplotype Analysis

Surveys of the *DMPK* CTG repeat size on normal chromosomes from global populations have shown a trimodal distribution of (CTG)_5_, (CTG)_8-17_, (CTG)_18-35_ repeat size ranges. These allelic bins correspond to three common haplotypes containing the *DMPK* CTG repeat within a strong block of linkage disequilibrium (LD), with most pathogenic CTG repeat expansions occurring on the (CTG)_18-35_ repeat haplotype.^5^ These haplotypes were defined by genotyping biallelic markers spanning a distance of 21 kb that included a 1-kb deletion of an *Alu* element, a *Hinf*I restriction site and a *Taq*I restriction site polymorphism. We SNP tagged this haplotype block by using three single nucleotide polymorphisms (SNPs) including the originally described *Hinf*I (rs16939) and *Taq*I polymorphisms (rs10415988) and a SNP (rs4802275) in complete LD with the *Alu* structural variant. We used genome-wide single nucleotide polymorphism (SNP) genotypes from non-DM1 control samples to identify individuals homozygous for the three common SNP tagged *DMPK* region haplotypes, with (---) designating the CTG_18-35_ haplotype, (+++) designating the CTG_8-17_ haplotype, and (-+-) designating the CTG_5_ haplotype, with their reference/alternate alleles indicated by (-/+) symbols. Genotype imputation and phasing for these samples has been previously described ^23^ and haplotype clustering was performed using the *Haplostrips* tool.^24^ TP-PCR with MCA and fragment analysis was performed on 45 non-DM1 samples homozygous for each of the three common *DMPK* haplotypes to confirm the CTG repeat sizes.

## 3 RESULTS

Our three primer TP-PCR design amplifies a minimally sized 73 bp fragment from the 5’ end of the common *DMPK* (CTG)_5_ allele (**Figure 1A)** and 160 bp fragment from the 3’ TP-PCR. To demonstrate that MCA with saturating intercalating dye can discriminate CTG repeat size classes, we compared melt profiles from TP-PCR from both 5’ and 3’ ends of the *DMPK* (CTG)_n_ repeat. Unimodal melting behavior was observed for the smallest CTG repeat control (5::5 homozygotes), while for heterozygous control samples with a range of normal, intermediate, premutation, and fully expanded CTG repeats, the melting behavior shifted from unimodal to multimodal melt curves as the repeat length increased (**Figure 1B**). The 5’ and 3’ TP-PCR were comparable across a wide spectrum of CTG repeat sizes although the smaller 5’ TP-PCR amplicon enhanced resolution at lower temperatures; therefore, subsequent analyses included only the 5’ TP-PCR. To compare the melt profile using our protocol to a predicted melt profile based on the observed sizes and ratios of TP-PCR amplicons, we used primer extension reactions after TP-PCR to determine the actual fragment sizes and stoichiometries produced by these conditions from heterozygous samples with a normal CTG repeat (5::13) versus an expanded CTG repeat (5::480) **(Figure 2A**). We used thermodynamic calculations for the individual primer extension products to predict a curve (-dH/dT) for the negative derivative of helicity versus temperature ^21^ based on the fragment size of the primer extension peaks. We then weighted each individual curve by their peak heights observed in the primer extension and calculated a composite melt curve by summing these weighted curves (**Figure 2B**). The predicted -dH/dT curve was a fair approximation of the actual -dF/dT melt curve (**Figure 2C**), confirming that the shape of the -dF/dT melt curve between 83 °C and 95 °C is a composite of the underlying melting behavior of the TP-PCR fragment ladders and contains useful information about the CTG repeat size.

**Figure 2.**
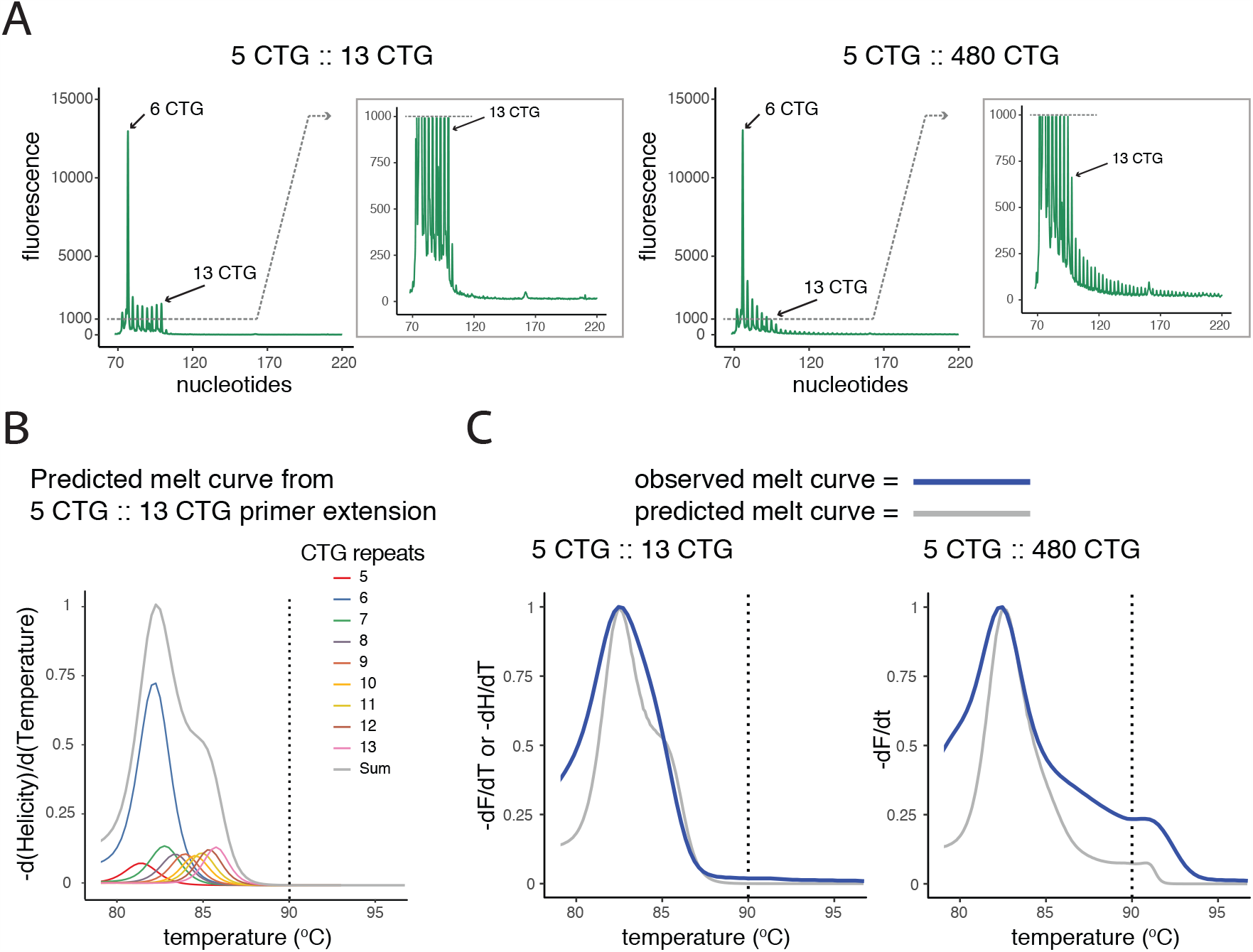
*DMPK* TP-PCR melt curve reconstruction. (A) Electropherograms from primer extensions using a FAM-labeled *DMPK* forward primer with post-TP-PCR products from a normal (5::13) and affected (5::480) individual. For each electropherogram, the boxed inset shows a zoomed y-axis. (B) Predicted -dH/dT melt curve (gray) summarized from the individual curves (colored) in proportion to their peak heights observed in (B) for the 5::13 sample. (C) The observed -dF/dT melt curves (blue) plotted with the normalized -dH/dT predicted curves (gray) for the 5::13 and 5::480 samples.

Melt profiles for nine control samples with a broad range of CTG repeat sizes were then analyzed, including four in the normal/intermediate range (5::5, 5::13, 5::14, and 14::30), one in the premutation range (12::37), and four in the fully expanded range (5::75, 5::80, 5::480, and 14::2530). Samples with larger CTG repeat sizes had more area under the -dF/dT melt curve at higher temperature than those with normal or smaller repeat sizes, and their melt profiles paralleled their predicted curves from primer extension analysis (**Figure 3A**). To determine the optimal amount of template DNA, we performed our TP-PCR assay using a serial dilution of genomic DNA from 20 ng to 0.02 ng, representing a range of ∼6000 to 6 genome copy equivalents per reaction. Our results indicated that the TP-PCR assay and MCA were robust across a broad range of DNA concentrations, with successful amplification in as low as 0.02 ng genomic DNA (**Figure 3B)**. Ultimately, we used ∼2 ng template for all other analyses.

**Figure 3.**
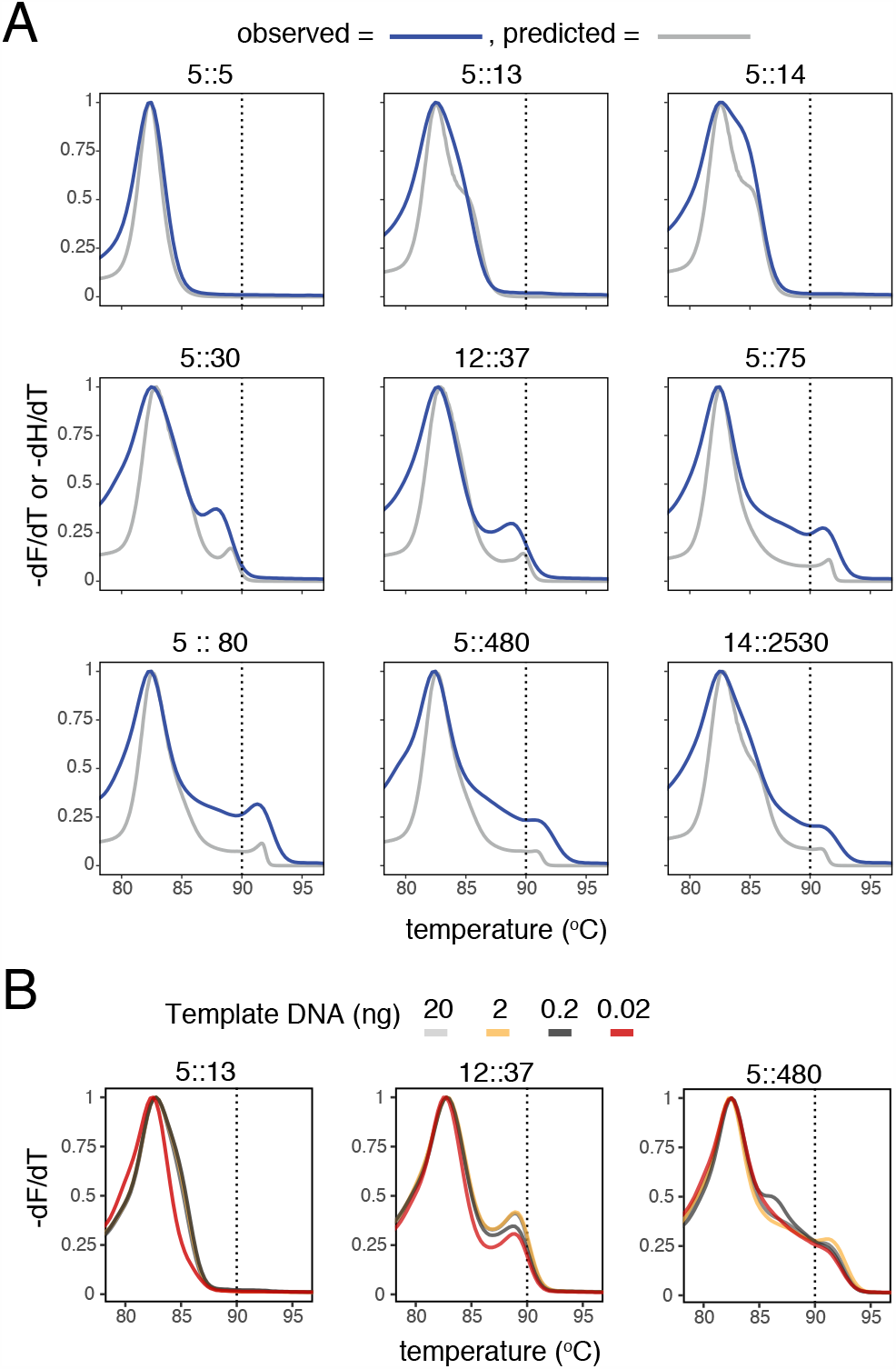
-dF/dT melt curves from normal control and DM1 patient samples. (A) Normalized -dF/dT melt curves from known CTG repeat lengths (blue) plotted against their predicted -dH/dT curves (gray). TP-PCR reactions used 2.0 ng of genomic DNA and the predicted melt curve were reconstructed from the fragment ratios observed by primer extension with post-TP-PCR products. (B) -dF/dT melt curves produced by TP-PCR using serially diluted amounts of human genomic DNA.

To better understand the behavior of the melt profile on control samples from the normal population, we chose 45 individuals who were homozygous for each of the three common *DMPK* haplotype groups for further analysis (**Figure 4A**). We sized the CTG repeats from these 45 individuals and observed a similar trimodal distribution of repeat sizes that recapitulated the original association of CTG_5_, CTG_8-17_, and CTG_18-35_ size ranges associated with the *Alu*/*Hinf*I/*Taq*I haplotypes in worldwide populations ^5, 23^. The (`-+-`)/(`-+-`) diplotype had exclusively CTG_5_ repeats, the (`+++`)/(`+++`) diplotype had repeat sizes in the CTG_8-14_ range, and the (`---`)/(`---`) diplotype had repeat sizes in the CTG_14-40_ range (**Figure 4B**). We screened these 45 homozygous *DMPK* diplotype samples and 57 individuals with clinical features of DM1 using the TP-PCR assay with MCA and observed unimodal melt profiles from the (`-+-`)/(`-+-`) and (`+++`)/(`+++`) diplotype groups, and multimodal melt profiles from the (`---`)/(`---`) diplotype and clinically affected DM1 subjects (**Figure 4C**). Individual melt profiles for each sample were scored by blinded reviewers as normal (unimodal below 90 _o_C), intermediate (bimodal below 90 °C), premutation (bimodal crossing 90 °C), or expanded (multimodal above 90 °C) and a classification was made based on the consensus of the blinded calls as detailed in Methods. All 57 DM1 subjects were classified as expanded by blinded review, while all the (`-+-`)/(`-+-`) and (`+++`)/(`+++`) diplotype samples were classified as normal (**Table 1**). The 15 (`---`)/(`---`) diplotype samples showed more variation, with blinded scores in three out of the four classes. The two samples that scored as normal had the smallest sizes (14 and 17 repeats), the nine samples scored as intermediate had intermediate sizes (21 to 28 repeats) and the four samples scored as premutation had the largest sizes (29 to 40 repeats) (**Supplemental Table 1**).

**Table 1.** Summary of blinded scoring of melt profiles from control samples with homozygous *DMPK* diplotypes and known DM1 patient samples.

|  | Blinded Score<sup>†</sup> |  |  |  | Total |
| --- | --- | --- | --- | --- | --- |
|  | Normal | Intermediate | Premutation | Expanded |  |
| (-+-) diplotype | 15 (100%) | - | - | - | 15 |
| (+++ ) diplotype | 15 (100%) | - | - | - | 15 |
| (---) diplotype | 2 (13%) | 9 (60%) | 4 (27%) |  | 15 |
| Known DM1 | - | - | - | 57 (100%) | 57 |

<sup>†</sup>Expected range for (CTG)<sub>n</sub> in the (-+-) haplotype=5, in the (+++) haplotype= 8-17 and in the (---) haplotype = 18-35.

**Figure 4.**
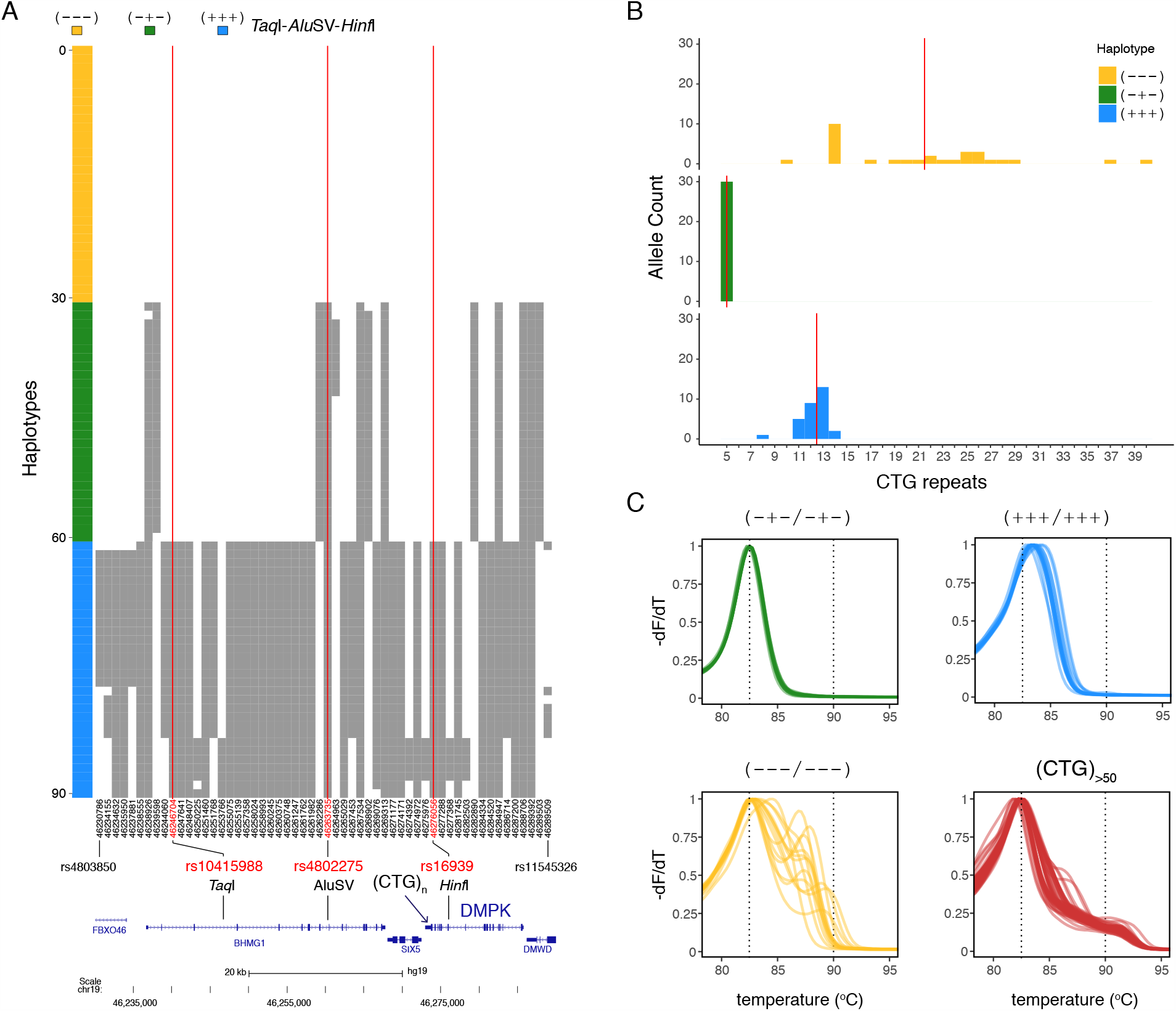
Haplotype and Melt analysis of common *DMPK* diplotypes and known DM1 patient samples. (A) *Haplostrips* plot of the *DMPK* region (chr19: 46230001-46290000, hg19) using 90 sorted and clustered haplotypes from 45 control samples. Phased haplotypes are in rows and SNPs with a minor allele frequency ≥ 0.15 are in columns. The alleles are colored relative to the (`---`) haplotype group with white as the reference allele and grey as the alternate allele for each of the 56 polymorphic sites. Vertical red lines indicate the positions of the original *Taq*I (rs10415988) and *Hinf*I (rs16939) polymorphisms, as well as a SNP (rs4802275) in complete LD with the *Alu* structural variant. (B) Plot of CTG repeat numbers observed for the 15 control samples within each of the three homozygous diplotype groups. Red vertical line indicates the median CTG repeat number in each diplotype group. Expected range for (CTG)_n_ in the (`-+-`) haplotype=5, in the (`+++`) haplotype= 8-17 and in the (`---`) haplotype = 18-35. (C) -dF/dT melt curves from 15 individual samples in each of the three common diplotype groups seen in human populations (green (`-+-`), blue (`+++`), and yellow (`---`), see Supplemental Table S1. In each group, diplotype colors and labels are the same as in (B), and melt curves from 57 clinically diagnosed DM1 patients are plotted in red.

To demonstrate use of this assay on a larger scale, applicable to population level studies, we performed our 5’ TP-PCR assay on 10,224 de-identified samples derived from DBS along with 522 control samples (58 replicates of 9 samples with known repeat sizes). Melt profiles of controls and DBS samples were reviewed by 4 blinded reviewers and scored as normal, intermediate, premutation, expanded, uncertain, or failed based on visual assessment of the melt as described above. Replicate control samples were scored by blinded reviewers and in most cases were classified correctly based on the known allele size (**Table 2**). Positive controls, with CTG repeats ≥50, were classified as expanded in all cases. The one premutation control, with 37 CTG repeats, was classified as premutation in 57 of 58 replicates. The control sample in the intermediate range (30 CTG repeats) showed more variability in the blinded scoring. In this case, 34 (58.6%) were scored as intermediate, 23 (39.6%) as premutation, and 1 (1.7%) as expanded. For the three control samples in the normal range (5-14 CTG repeats), 93-96% of these replicates were scored as normal.

**Table 2.**
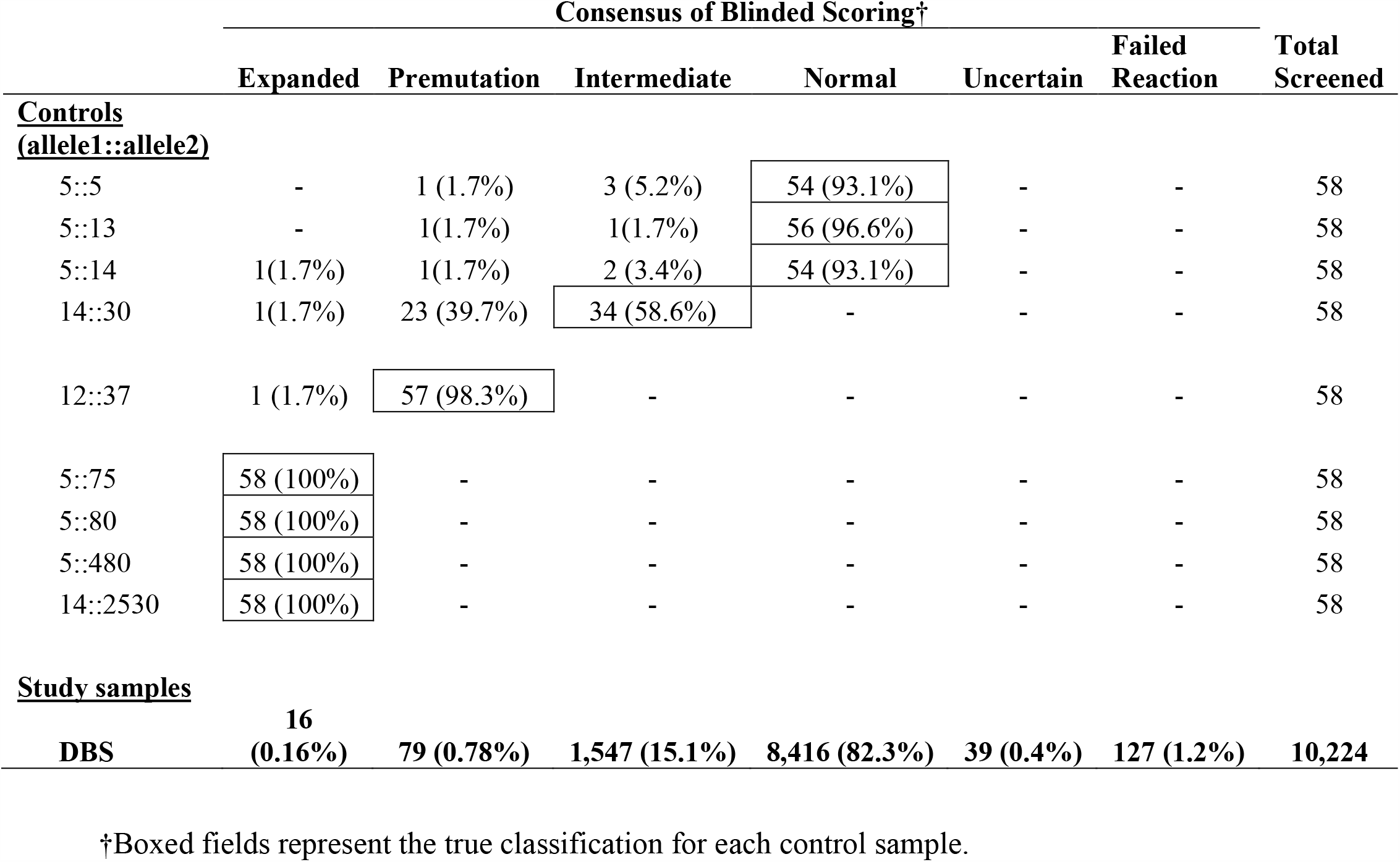
Summary of blinded scoring from control samples and dried blood spots.

For the 10,224 DBS samples, we obtained a successful melt profile in 10,097 (98.76%) (**Table 2**). From these 8,416 (82.3%) were scored by blinded reviewers as normal and 1,547 (15.1%) as intermediate, in good agreement with observed frequencies of the CTG_5_ and CTG_8-17_ versus the CTG_18-35_ haplotypes in non-African populations ^5^. We determined CTG repeat sizes from 142 intermediate samples and observed that 121 (85%) had a repeat size between 18 and 30 (median size = CTG_24_) for the larger allele, indicating that the intermediate class was enriched for the CTG_18-35_ (`---`) haplotype. The remaining 21 samples were in the normal range with size <18 repeats. For an unbiased evaluation of the melt curve profiles based on the full set of 10,097 samples, we used the -dF/dT melt curves in a clustering analysis to construct a UMAP visualization. The clustering analysis demonstrated proximity and clustering of similar size repeats (**Figure 5A**).

**Figure 5.**
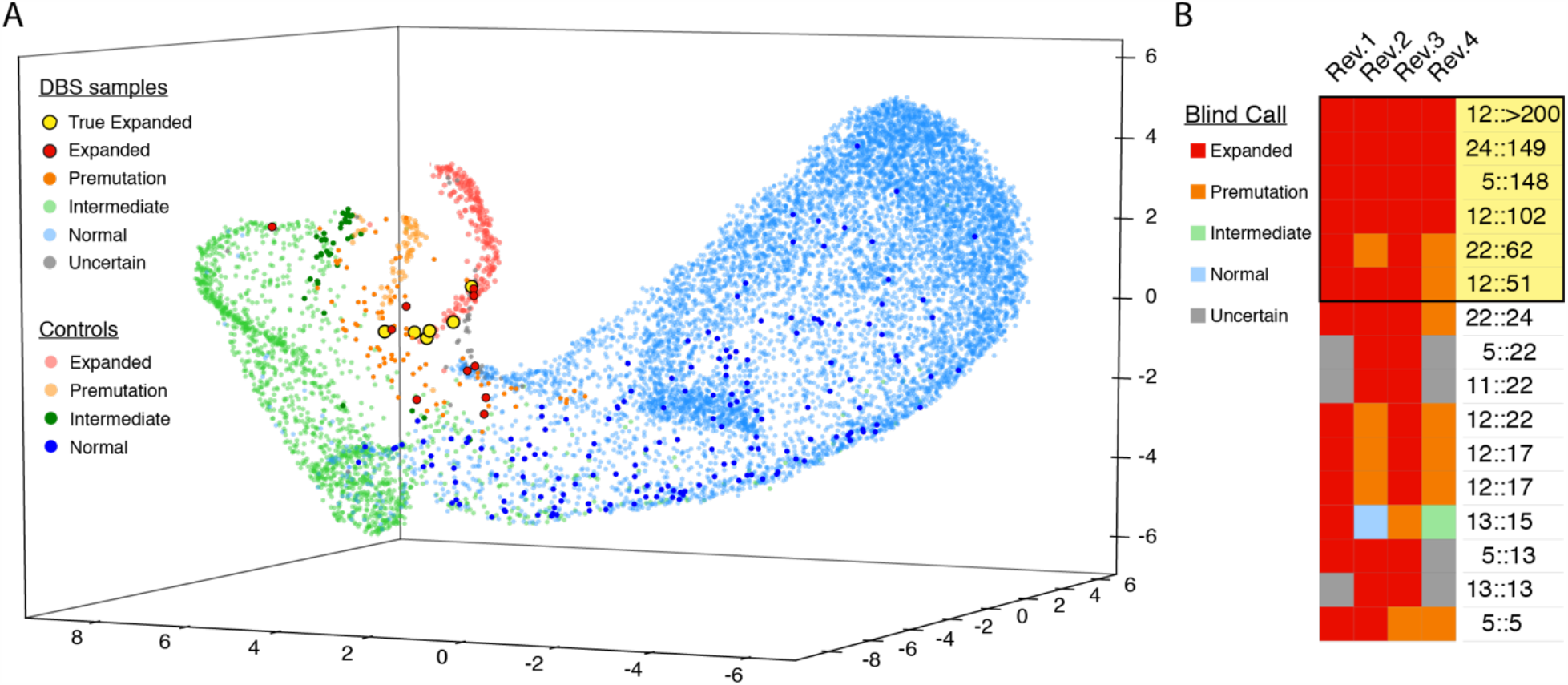
*UMAP* clustering and variability of blinded review of melt curves from DBS. (A) The -dF/dT data from 87.25 °C to 96.70 °C was clustered by UMAP and visualized as a 3-dimensional representation. 10,097 DBS samples, and 232 expanded (5::75, 5::80, 5::480, and 5::2530), 56 premutation (12::37), 44 intermediate (14::30), and 170 normal (5::5, 5::13, and 5::14) control melt profiles were used with sample points colored by the consensus call of blinded reviewers. (B) Heatmap of individual calls from the four blinded reviewers and the final sized CTG repeat size (allele 1 :: allele 2) by direct PCR or the Amplidex® DM1 Dx Kit for each sample with consensus call of ‘expanded’’ is shown, with the true positives boxed in yellow.

The blinded review of 10,097 DBS samples resulted in 16 samples (0.16%) classified as expanded (**Figure 5B, Supplemental Figure 2**) and 79 as premutation (0.78%) (**Supplemental Figure 3**). All samples classified as expanded or premutation by blinded review were referred for direct sizing performed by fluorescent PCR across the CTG repeat or using the Amplidex DM1 Dx Kit. Six of the 16 samples identified by screening were found to have CTG repeats ≥50, with expanded allele sizes of 51, 62, 102, 148, 149, and >200 CTG repeats (**Figure 6**). Six of the samples that screened as false positive had allele sizes in the intermediate range (22::24, 5::22, 11::22, 12::22, 12::17, 12::17) and four had allele sizes in the normal range (13::15, 5::13, 13::13, 5::5). For the 79 samples that were classified as premutation by blinded reviewers, 16 were found to have true CTG expansions in the premutation range with allele sizes from 35-49, 26 were found to have CTG repeat alleles in the intermediate range, and 37 with CTG repeat alleles in the normal range (**Supplemental Figure 3**). None had full expansion of the CTG repeat (≥ 50).

**Figure 6.**
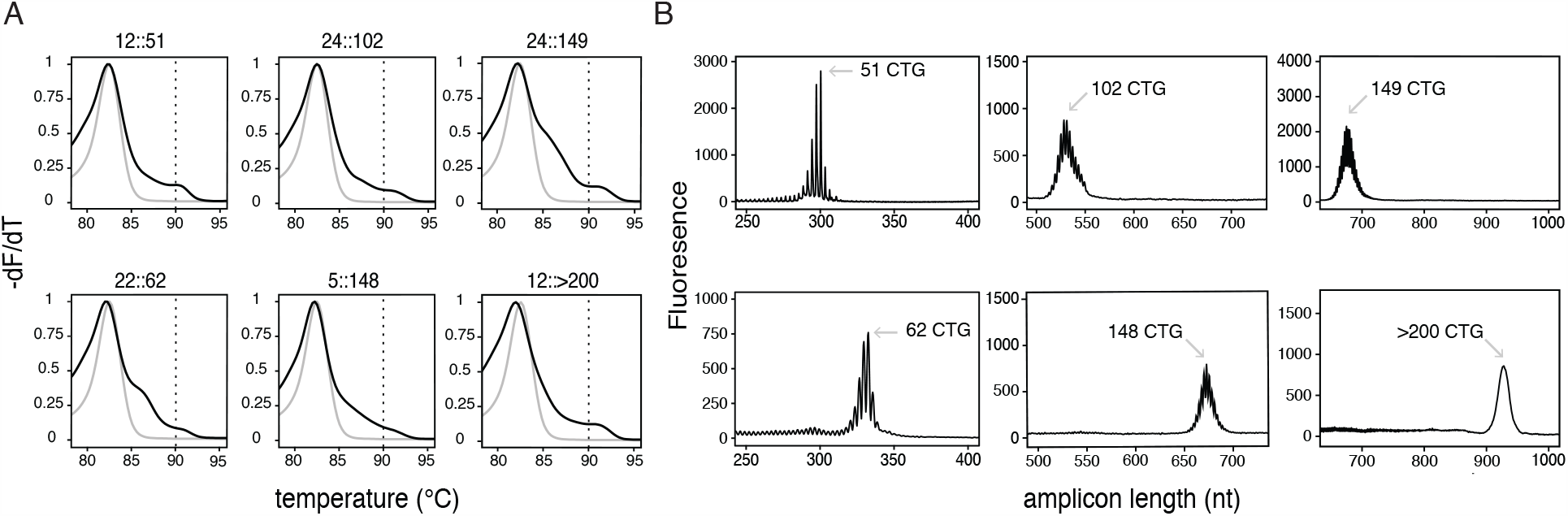
Melt curve profiles and sizes for DBS samples with CTG repeats ≥50. (A) Normalized -dF/dT melt curves for the 6 size-confirmed expanded samples (black), plotted with normal (5::5) control (gray). (B) Capillary electropherograms of the 6 expanded DBS samples amplified with direct sizing PCR (51 and 62 CTG repeats) or using the Amplidex® DM1 Dx Kit (102, 148, 149 and >200å CTG repeats).

## 4 DISCUSSION

Here we present a low-cost, high throughput assay for identifying expanded CTG repeats in the *DMPK* gene in a single tube using TP-PCR with MCA with the saturating dye SYBR GreenER. This assay relies on identifying a shift in melting profile in samples with expanded CTG repeats that is readily identified by visual examination of the melt profile. Using this assay, we were able to correctly detect expansions ≥ 50 CTG repeats with 100% sensitivity (232 of 232 control samples classified as expanded) and 99% specificity (287 of 290 control samples classified not expanded). In addition, our assay is able to detect shifts in the melt profile from samples with CTG repeats in the premutation (35-49 repeats) and intermediate (18-34 repeats) ranges but with less sensitivity/specificity. Our ability to identify samples with the putative (CTG)_18-35_ risk haplotype as intermediate melt profiles observed in 15% of our samples confirms results from earlier *DMPK* haplotype analysis in worldwide populations and indicates the utility of using a full spectrum of fragment lengths in the melt analysis.

Our assay is robust for use with 1-2 ng template DNA from a simple DNA preparation on DBS samples, enhancing scalability for large screens. While we use 1-2 ng of template DNA for our study here, our assay is successful with as low as 0.02 ng (6 genome copy equivalents) input DNA, making the assay practical for use with samples derived from sources with limited DNA. In contrast, alternative methods using for TP-PCR with MCA for the CTG repeat in *DMPK* require as much as 100 ng input DNA and lose signal below 5ng.^25^

Our assay builds on previous methods to use MCA to detect repeat expansion alleles. Multimodal melt curves are typically observed in MCA with mixtures of PCR amplicons of different lengths or sequence, since intercalating dyes bind non-specifically to any double-stranded DNA molecules. An alternative TP-PCR/MCA method uses non-saturating conditions with SYBR Green I dye, where the limiting dye dissociates from melting lower T_m_ molecules and re-intercalates to higher T_m_ double-strand molecules during a slow temperature ramp, resulting in a unimodal derivative melt peak observed only at the highest melting temperature.^16, 25^ While this elegant assay simplifies the melt curve interpretation, it requires precise matching of dye concentration with the final concentration of PCR amplicons that may be difficult with lower quality DNA samples.

Notably, a similar TP-PCR/MCA assay has been successfully used for screening DBS samples for triplet repeat expansion in fragile X syndrome, but that assay requires significantly more template DNA (a minimum of 20ng DNA vs 1-2ng DNA for our assay) from a more complex DNA purification using a column prep.^26^ While successful in screening, it may be difficult to scale up this process to thousands of samples. In our study, we re-evaluate the premise that non-saturating conditions in TP-PCR with MCA is superior to saturating conditions. We show that the use of MCA with the saturating SYBR GreenER dye has comparable resolution in distinguishing the relevant range of CTG repeat expansions. In our assay, we use SYBR GreenER dye for the high resolution melt rather than other dyes commonly used in high resolution melt applications. While it is likely that other saturating dyes such as (LC Green 1, LC Green +, or EvaGreen) are also suitable for this purpose, we favor the use of SYBR GreenER because is available in a commercially available master mix (SYBR Select) which adds simplicity to the reaction preparation and scalability and minimizes variability from the reagent preparation.

To demonstrate the feasibility of this assay for deployment in a large-scale population screen, we tested 10,224 DBS samples. We successfully screened 10,097 samples, with a reaction failure rate of only 1.2%. Sixteen samples were identified as expanded from screening by blinded reviewers based on MCA and secondary validation steps revealed true CTG expansions ≥ 50 in 6 of these samples. The ten false positive samples had CTG repeat sizes in the normal to intermediate range (5 to 24 CTG repeats), resulting in positive predicted value (PPV) of 37.5% based solely on the TP-PCR/MCA which compares favorably with screening protocols for other disorders such as cystic fibrosis.^27^ For a combined screening approach with TP-PCR/MCA followed by the more definitive test, all false positive samples were eliminated, reflecting a PPV approaching 100% for the two-step process. For 79 samples classified as premutation by MCA, 16 were found to have true CTG repeats in the premutation range (35-49 repeats) resulting in positive predicted value of 20.3% to detect premutation carriers based solely on the TP-PCR/MCA. While we are not able to determine negative predictive value since we cannot fully exclude false negative samples, we presume that the actual number of false negatives is very low based on the absence of false positive cases in our control samples (Tables 1 and 2) and the higher than expected number of true positive samples identified here (6 per 10,000) compared to published prevalence (0.5-1.5 per 10,000).^6-8^ Notably, in no case was a sample that screened as intermediate or premutation by blinded review of the MCA found to have an actual repeat larger than its initial classification.

While our method adds technical simplicity to the TP-PCR with MCA method since we use saturating rather than non-saturating conditions, it has several limitations. The melt profile from our assay incorporates information from a mixture of TP-PCR products of different lengths from different alleles so the melt profile can have some heterogeneity. Interpretation of the melt profile by visual inspection can be subjective and depends somewhat on the skill experience of the reviewer. While our study was focused on assay development and included primarily visual scoring of the melt profile, our clustering analysis suggests that automated classification of melt profiles is feasible for screening of larger populations (Figure 5). While we can identify samples with suspected expanded CTG repeats ≥50, and repeats in the premutation and intermediate range, we cannot determine the exact size of the repeat without follow-up testing by direct PCR or fluorescent TP-PCR, with a limit of resolution of ∼200 repeats by capillary electrophoresis. For large-scale screening purposes, our assay is highly sensitive, but does result in some false positive results (10 false positive samples in 10,224 samples tested) which require follow-up testing for confirmation, however, the TP-PCR/MCA vastly narrows the search and limits the number of samples that require this type of validation. For false positive samples, we suspect that the initial TP-PCR/MCA assays may have had non-specific PCR amplicons that contributed to the melt profile. This suggests that the specificity of the initial TP-PCR primer design and PCR conditions are an important aspect for developing a robust assay.

An emerging limitation to the wider application of this technique is the competing method of repeat expansion profiling using next generation sequencing. Recent application of whole genome sequencing using short Illumina reads (100 – 150 bases) detected 15 *DMPK* expansions in 12,632 genomes. That study included several DM1 families that may have contributed to the higher number of detected expansions but is methodologically a powerful alternative to TP-PCR MCA assays ^28^. Still, at an estimated cost of $0.89 per sample for a screen that can be done in a single tube, our method is less costly and complex than next-generation sequencing strategies.

While we have optimized our TP-PCR with MCA from both 5’ and 3’ ends of the (CTG)_n_ expansion in *DMPK*, we tested only the 5’ assay in the larger cohort of DBS samples. Past recommendations to include both 5’ and 3’ TP-PCR stem from the possibility that expanded samples may be missed in TP-PCR reactions due to interruptions of the CTG repeat.^11, 16, 29, 30^ However, bidirectional assays are likely unnecessary, particularly in the setting of a screening test as we have proposed here. With traditional TP-PCR careful examination of the electropherograms from published reports of DM1 cases with interrupted CTG repeats show that the TP-PCR reaction does not fail, but proceeds with interruptions to the typical sawtooth pattern seen from uninterrupted CTG repeats.^11, 29, 31, 32^ Indeed, TP-PCR has been used in search of interrupted repeats.^33^ As we have shown by melt curve reconstruction, since our application of MCA uses the full spectrum of TP-PCR products, it is robust at detecting differences in the relative proportion of fragment lengths. Furthermore, interruptions in the CTG repeat in *DMPK* are rare (3-5% of cases) and almost always occur in the 3’ end of the CTG repeat and would be least disruptive to the pattern of TP-PCR amplicons in our assay.^29, 31, 34^

DM1 is one of the most prevalent muscular dystrophies and diagnostic delays as long as seven years are common.^9^ Next-generation sequencing panels are increasingly used as the first diagnostic test for patients with suspected neuromuscular disorders,^35^ but will miss disorders due to triplet-repeat expansions such as DM1. Our assay can easily be incorporated into diagnostic panels greatly facilitating early diagnosis and avoiding additional unnecessary testing. The low cost and simplicity of this assay make it an ideal tool for screening programs interested in identifying both patients with DM1 and also patients with premutation alleles at risk for passing expanded CTG repeat alleles to children. Further, this technique can be easily adapted to population screens for other disorders caused by repeat expansions such as type 2 myotonic dystrophy, Huntington disease, Fragile X syndrome, and others.

## Data Availability

The data that support the findings of this study are available from the corresponding author upon reasonable request.

## ACKNOWLEDGEMENTS

We are grateful to the Myotonic Dystrophy Foundation for funding for this study. Additional funding from K08 NS097631 (RJB), R21 NS100040 (RBW), K23NS091511 (NEJ). We are grateful to Kaycie Lawson, MS, Mark Shook, BS, Zoe Edmunds, BS, Denise Kay, PhD and Michele Caggana, ScD, FACMG of the New York State Newborn Screening Program, Wadsworth Center, New York State Department of Health for providing deidentified samples, and to Michael Klein, BS of the University of Utah Genomics Core for providing instrumentation and expertise for acquiring melt curves.

