## Supplemental Table and Figures for "High Throughput Screening for Expanded CTG repeats in Myotonic Dystrophy Type 1 Using Melt Curve Analysis"

**Supplemental Figure 1. Template for scoring TP-PCR melt curves.**
Four examples of typical melt curves as reviewed by the blinded reviewers.  Each sample classified into one of six categories based on visual inspection of the MCP: Normal, Intermediate, Premutation, Expanded, Uncertain, and Fail. The melt curve for the DBS samples (black) is plotted with averaged melt curves from the four positive (expanded) controls with $(CTG)_n \geq 50$ [5::75 (green), 5::80 (purple), 5::480 (light red), and 14:2530 (light blue)].

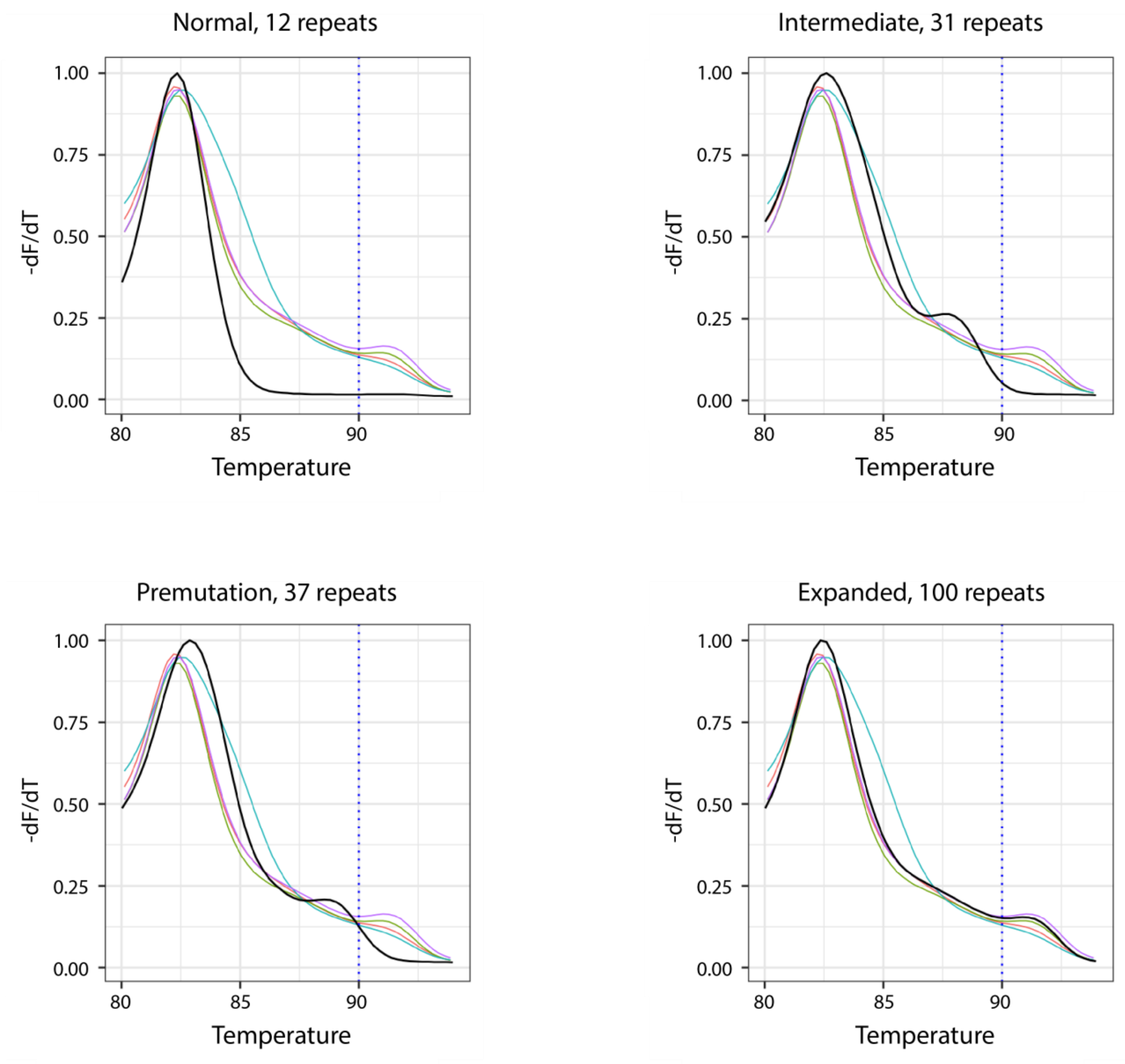

**Supplemental Figure 2. Melt curve profiles from all samples classified as 'expanded' by blinded scoring.** Each sample was reviewed by 4 blinded reviewers, with 16 samples resulting in a 'expanded' score based on the consensus of the reviewers. The melt curve for the DBS sample (black) is plotted with averaged melt curves from four positive (expanded) controls with $(CTG)_n$ ≥50: 5::75 (green), 5::80 (purple), 5::480 (light red), and 14:2530 (light blue). Repeat numbers for Allele 1:: Allele 2 by direct PCR or the Amplidex® DM1 Dx Kit are shown above each plot. Samples with true expansions are highlighted in yellow.

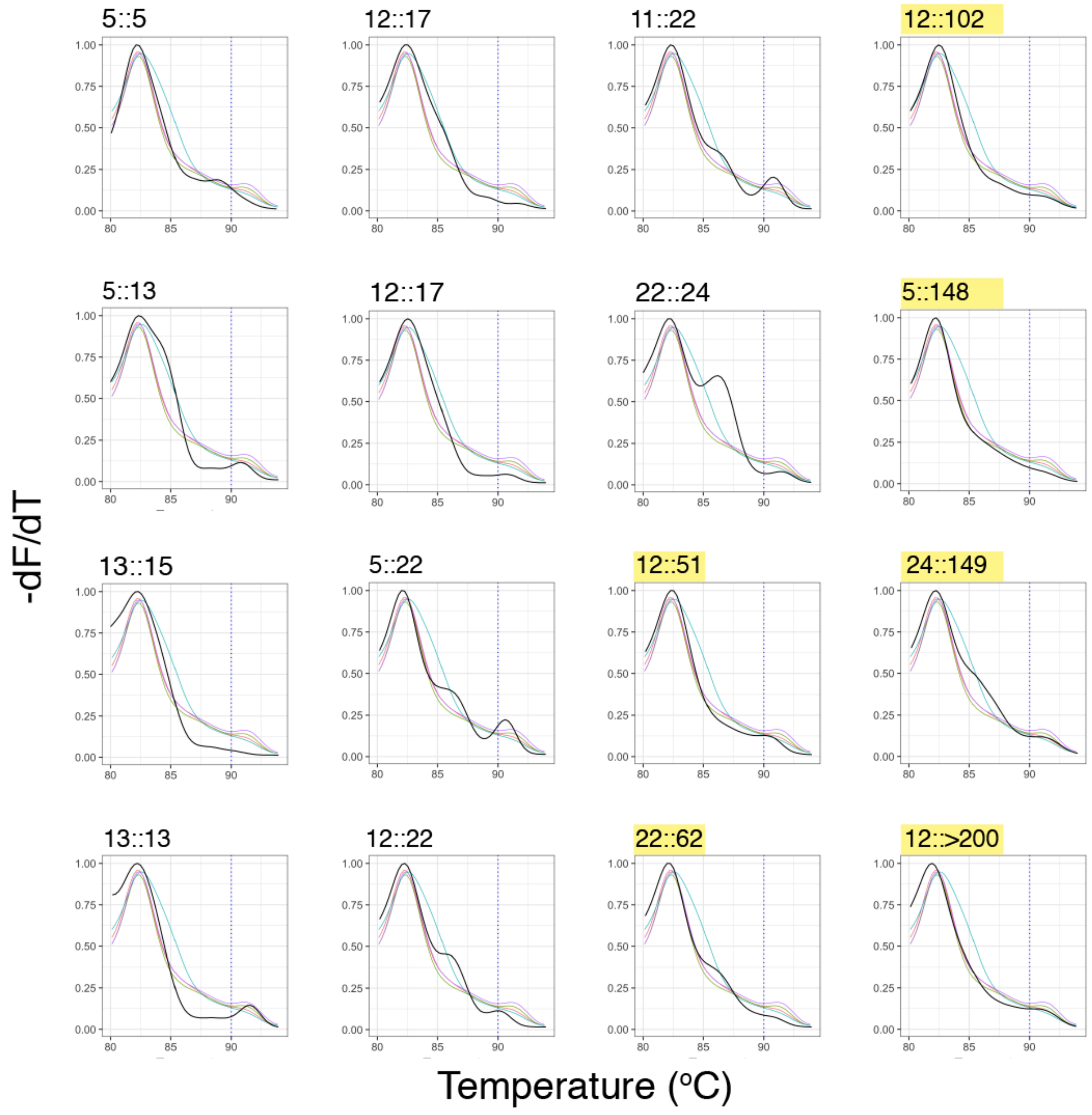

**Supplemental Figure 3. Individual scores of melt curve profiles and validated CTG repeat size from all samples classified as 'premutation' by blinded scoring.** Heatmap of individual calls from the four blinded reviewers and final CTG repeat numbers for Allele 1 :: Allele 2 by direct PCR or the Amplidex® DM1 Dx Kit are shown for each sample, with the true premutations boxed in yellow.

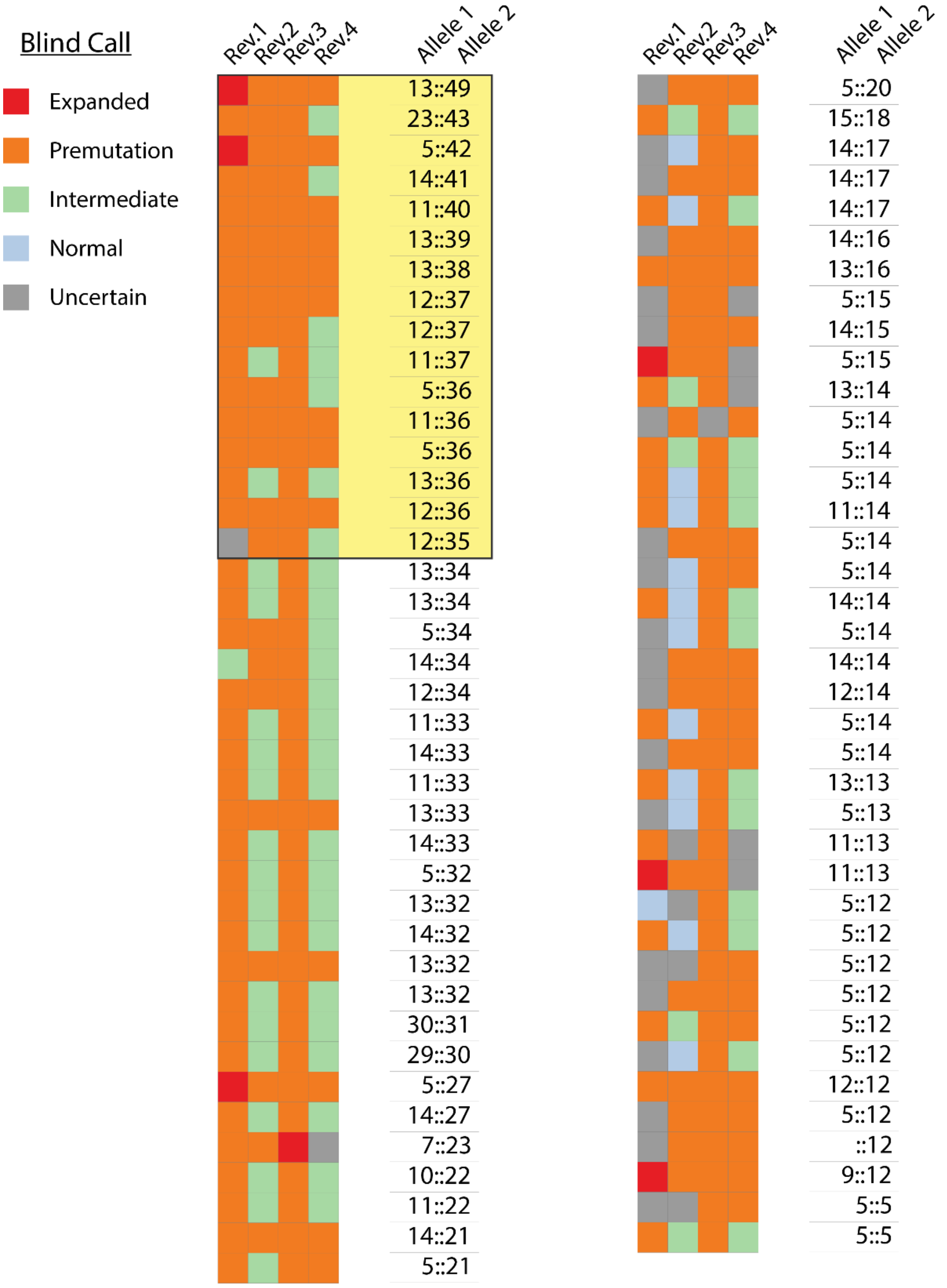

**Supplemental Table 1. Individual allele sizes and blinded scores from for control samples.**

| Sample ID | Diplotype Group† | Allele 1 $(CTG)_n$ | Allele 2 $(CTG)_n$ | Consensus Score from Blinded Review |
|---|---|---|---|---|
| 1 | (-+-) | 5 | 5 | Normal |
| 2 | (-+-) | 5 | 5 | Normal |
| 3 | (-+-) | 5 | 5 | Normal |
| 4 | (-+-) | 5 | 5 | Normal |
| 5 | (-+-) | 5 | 5 | Normal |
| 6 | (-+-) | 5 | 5 | Normal |
| 7 | (-+-) | 5 | 5 | Normal |
| 8 | (-+-) | 5 | 5 | Normal |
| 9 | (-+-) | 5 | 5 | Normal |
| 10 | (-+-) | 5 | 5 | Normal |
| 11 | (-+-) | 5 | 5 | Normal |
| 12 | (-+-) | 5 | 5 | Normal |
| 13 | (-+-) | 5 | 5 | Normal |
| 14 | (-+-) | 5 | 5 | Normal |
| 15 | (-+-) | 5 | 5 | Normal |
| 16 | (+++) | 8 | 13 | Normal |
| 17 | (+++) | 11 | 11 | Normal |
| 18 | (+++) | 12 | 12 | Normal |
| 19 | (+++) | 12 | 12 | Normal |
| 20 | (+++) | 13 | 13 | Normal |
| 21 | (+++) | 11 | 13 | Normal |
| 22 | (+++) | 12 | 13 | Normal |
| 23 | (+++) | 13 | 13 | Normal |
| 24 | (+++) | 12 | 13 | Normal |
| 25 | (+++) | 11 | 13 | Normal |
| 26 | (+++) | 12 | 13 | Normal |
| 27 | (+++) | 12 | 13 | Normal |
| 28 | (+++) | 12 | 13 | Normal |
| 29 | (+++) | 11 | 13 | Normal |
| 30 | (+++) | 14 | 14 | Normal |
| 31 | (---) | 14 | 14 | Normal |
| 32 | (---) | 14 | 17 | Normal |
| 33 | (---) | 14 | 21 | Intermediate |
| 34 | (---) | 14 | 22 | Intermediate |
| 35 | (---) | 22 | 23 | Intermediate |
| 36 | (---) | 14 | 25 | Intermediate |
| 37 | (---) | 14 | 25 | Intermediate |

| | | | | |
|---|---|---|---|---|
| 38 | (---) | 10 | 25 | Intermediate |
| 39 | (---) | 14 | 26 | Intermediate |
| 40 | (---) | 24 | 26 | Intermediate |
| 41 | (---) | 27 | 28 | Intermediate |
| 42 | (---) | 26 | 29 | Premutation |
| 43 | (---) | 12 | 37 | Premutation |
| 44 | (---) | 14 | 37 | Premutation |
| 45 | (---) | 14 | 40 | Premutation |

† Expected range for $(CTG)_n$ in the (-+-) haplotype=5, in the (+++) haplotype= 8-17 and in the (---) haplotype = 18-35.
